# The more symptoms the better? Covid-19 vaccine side effects and long-term neutralizing antibody response

**DOI:** 10.1101/2023.09.26.23296186

**Authors:** Ethan G. Dutcher, Elissa S. Epel, Ashley E. Mason, Frederick M. Hecht, James E. Robinson, Stacy S. Drury, Aric A. Prather

## Abstract

Protection against SARS-CoV-2 wanes over time, and booster uptake has been low, in part because of concern about side effects. We examined the relationships between local and systemic symptoms, biometric changes, and neutralizing antibodies (nAB) after mRNA vaccination. Data were collected from adults (n = 364) who received two doses of either BNT162b2 or mRNA-1273. Serum nAB concentration was measured at 1 and 6 months post-vaccination. Daily symptom surveys were completed for six days starting on the day of each dose. Concurrently, objective biometric measurements, including skin temperature, heart rate, heart rate variability, and respiratory rate, were collected. We found that certain symptoms (chills, tiredness, feeling unwell, and headache) after the second dose were associated with increases in nAB at 1 and 6 months post-vaccination, to roughly 140-160% the level of individuals without each symptom. Each additional symptom predicted a 1.1-fold nAB increase. Greater increases in skin temperature and heart rate after the second dose predicted higher nAB levels at both time points, but skin temperature change was more predictive of durable (6 month) nAB response than of short-term (1 month) nAB response. In the context of low ongoing vaccine uptake, our convergent symptom and biometric findings suggest that public health messaging could seek to reframe systemic symptoms after vaccination as desirable.

## Introduction

Vaccination against SARS-CoV-2 has been repeatedly shown to reduce infections, hospitalizations, and mortality, but protection wanes considerably over time for all of these outcomes, even following booster vaccination (1). Moreover, uptake of booster vaccinations has been low, with only 17% of the US population having received the bivalent booster as of May 2023, despite the vaccine having been widely available for over six months at that time (2). Among individuals who received at least one dose of a COVID-19 vaccine, the most commonly reported reasons for not having received a booster were: first, a perception of low added benefit in protection from illness, given a personal history of prior vaccination or SARS-CoV-2 infection, and second, worry about side effects (3,4).

Recent evidence has suggested that greater systemic symptoms following SARS-CoV-2 vaccination may reflect a more potent immune response (5–7). A deeper understanding of this relationship may help to address low rates of vaccine uptake. If the association is clinically meaningful, public health messaging might aid uptake by reframing short-term post-vaccination symptoms as positive indications that the vaccine is likely to be working rather than undesirable side effects.

Although there are several reports suggesting that SARS-CoV-2 vaccine reactogenicity (i.e., resulting symptom burden or physiological perturbation) predicts higher subsequent anti-spike immunoglobulin level (5–7), only a small number of studies have specifically measured neutralizing antibodies (nAB) (8–10). Results from these studies are inconsistent, and they have only measured short-term nAB responses. Quantifying functional antibody activity (i.e., nAB) is important because although they are correlated, vaccine effects on nAB and absolute anti-spike IgG are dissociable, and nAB specifically appear critical in conferring protection from COVID-19. Only approximately 50% of the variability in nAB is predictable from anti-spike IgG (11), and nAB has been reported to have a larger effect size (i.e., lower hazard ratio per 10-fold increase) than anti-spike IgG in predicting subsequent COVID-19 incidence (12). It has been demonstrated that providing animals with neutralizing antibodies alone confers protection against disease even after high-dose SARS-CoV-2 exposure (13), and in one study in humans, nAB level was estimated to mediate over two-thirds of vaccine efficacy (12). A recent meta-analysis (14) and large pooled cohort analysis (15) of vaccination studies have estimated the correlation between average vaccine-evoked nAB and vaccine efficacy to be 0.81 and 0.91 respectively.

Using data from a cohort of adults who received the initial two-dose series of BNT126b2 or mRNA-1273, we used convergent self-report symptom and objective biometric measurements to identify predictors of subsequent serum nAB concentration at 1 and 6 months post-vaccination. For each vaccine dose, self-report variables included the presence or absence of 13 individual symptoms and total systemic symptom count. Biometric variables included measures of vaccination-induced change in skin temperature, heart rate, heart rate variability, and respiratory rate.

## Methods

### Subjects

Subjects were participants in the Building Optimal Antibodies Study (16), a large observational study designed to identify psychosocial, behavioral, and biological predictors of immune responses to COVID-19 vaccination. Participants were adults aged 18 years and above who did not have a previous history of immune-related disease and were not currently undergoing treatment with medications known to impact the immune system. Ethical approval was provided by the Institutional Review Board at the University of California, San Francisco, and all study participants provided written informed consent. The STROBE reporting checklist for cohort studies is provided in Supplement Table 1.

Serum was collected from study participants before they received a COVID-19 vaccine and again 1 and 6 months after they completed their initial two-dose series of BNT162b2 or mRNA-1273. Participants independently arranged to be vaccinated in the community, and vaccination date and type was later determined using official records. History of SARS-CoV-2 infection was examined by measuring levels of anti-spike IgG antibodies at baseline and anti-nucleocapsid IgG antibodies at 6 months. Participants with a positive result on either test were excluded from analyses. Ad26.COV2.S recipients were excluded from analyses given that use of this vaccine is no longer authorized by the US Food & Drug Administration. Other reasons for exclusion of subjects from analyses are provided in the flow chart in Figure 1.

**Figure 1.**
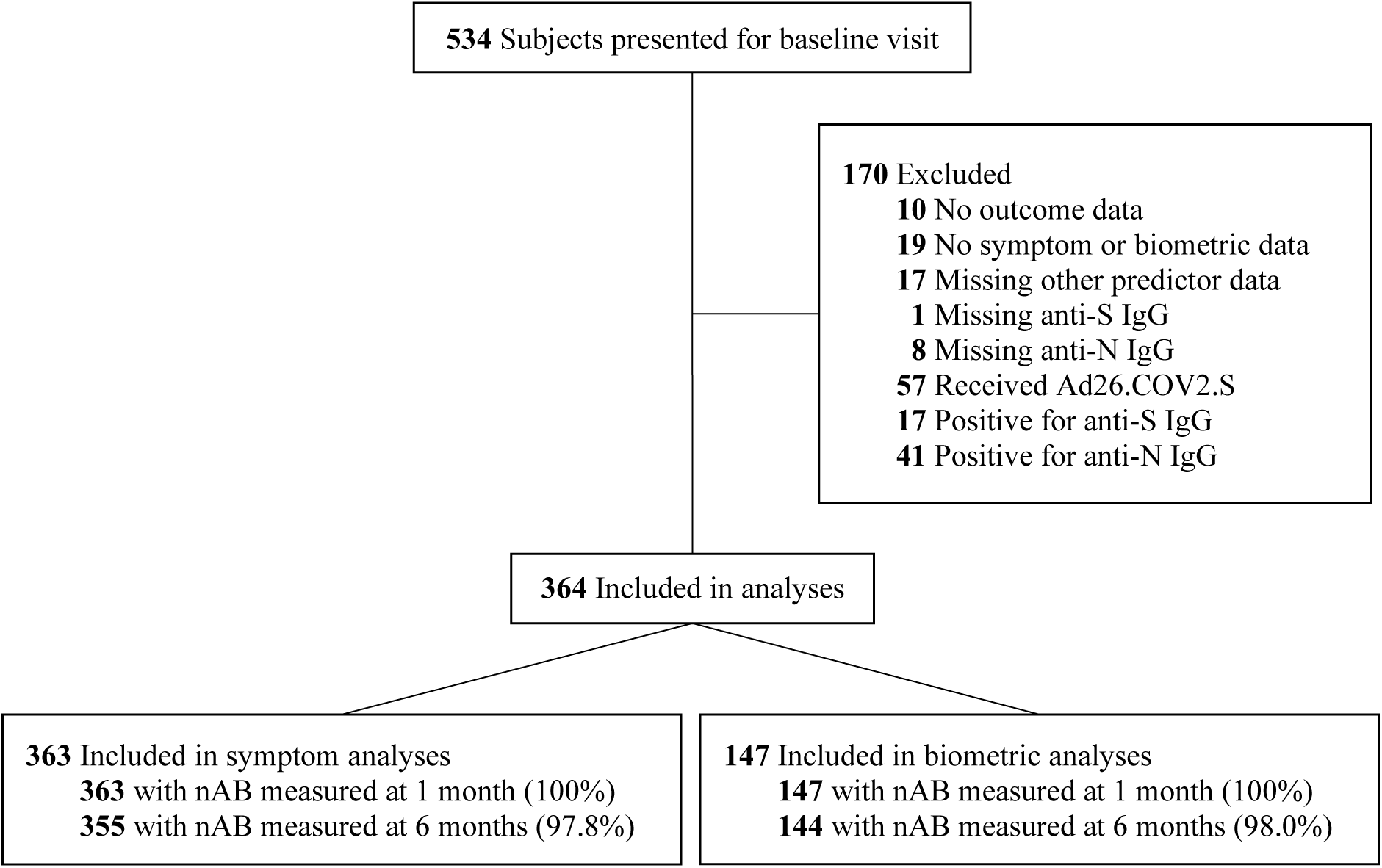
Flow chart of participants and observations. nAB = neutralizing antibodies; anti-S IgG = anti-spike immunoglobulin G; anti-N IgG = anti-nucleocapsid immunoglobulin G.

### Outcome

Neutralizing antibodies against SARS-CoV-2 were measured via pseudovirus assay at 1 and 6 months following vaccination as described previously.(16) In brief, serum from each participant was serially diluted and incubated with pseudovirus expressing full-length SARS-CoV-2 protein (Wuhan/D614G strain), permitting virion binding and neutralization by host antibodies. Serum-virus mixtures were then incubated with susceptible cells, allowed remaining functional pseudovirus to deliver a luciferase reporter gene intracellularly. After 66-72 hours, the medium was removed, lysis buffer and luciferase substrate were added, and luciferase activity was measured as luminescence. Neutralizing antibody titers were expressed as the inhibitory dose 50 (ID50), defined as the serum dilution corresponding to a reduction of relative light units (RLU) by 50% compared to serum-free control wells.

### Daily symptom surveys

Subjects were sent links to a survey each evening for six days, beginning on the date they anticipated receiving each dose of a SARS-CoV-2 vaccine. The survey included the question, “Did you experience any of the following physical symptoms today? (Check all that apply.)”. The following options were provided: Tiredness; Headache; Muscle pain; Chills; Joint pain; Fever; Nausea / vomiting; Feeling unwell; Tender or swollen lymph nodes (lymphadenopathy); Injection site pain, redness or swelling; Pain or swelling in the arm that did not get the vaccination; Other allergic reactions (difficulty breathing, swelling of face/throat, rash); Stomachache. For each survey entry, vaccine dose dates were used to calculate calendar days since receipt of either dose one or two. For each symptom, data were collapsed to reflect either presence on any of the six days or absence on all six days.

### Biometric collection and analysis

Heart rate (HR), heart rate variability (HRV), respiratory rate (RR), and skin temperature (ST) data were collected from a subset of participants using a biometric wearable device, the Oura Ring (Oura Health Ltd., San Francisco, CA, USA). Except for one individual, all individuals who provided biometric data were aged over 50 years, because only these individuals were actively offered devices.

During sleep, HR and HRV were recorded in 5-minute intervals while ST was recorded in 1-minute intervals. RR was only available as a nightly average. To test the hypotheses that short-term effects of vaccination on nighttime HR, HRV, ST, and RR are predictive of subsequent neutralizing antibody response, it was necessary to first derive a single summary value of vaccination-induced change in each physiological domain for each subject. For this purpose, for each domain, we used a multi-step procedure to identify the summarization approach with the best statistical evidence of vaccination-induced change (17,18); see the Supplement Methods for details. Ultimately, nightly time series of observations were first summarized into single nightly values for each subject: for ST and HRV by taking the nightly 99^th^ percentile (i.e., the “stable maximum”), and for HR by taking the nightly 1^st^ percentile (i.e., the “stable minimum”). Then, for each subject, for each physiological domain, the values on the first and second night following vaccination were each subtracted from a subject-specific norm. Finally, the greater of the two deviations from the subject norm (i.e., the vaccination-induced change on either the first or second night after vaccination), was taken as each subject’s single value of vaccination-induced change. Descriptive and test statistics for candidate summary variables of vaccination-induced change are provided in Supplement Table 2. Spearman correlations between final summary variables and symptom count are presented in Supplement Figure 1.

### Data analysis

All data analysis was performed in R v4.2.2. For all analyses, mixed-effects models were fit to nAB data collected at 1 month and 6 months following completion of the second vaccine dose. All models included a core set of terms, including a time point × vaccine interaction and main effects of vaccine, time point, sex, age, baseline smoking status, and body mass index. The statistical significance of these terms has been previously described (16). Here, for each vaccine dose, 18 variables were examined as predictors of subsequent nAB level: the presence or absence of 13 symptoms, the total count of reported symptoms (excluding injection site symptoms), and the levels of 4 biometric measurements. For each variable, a model was created by adding to the core model structure the following terms: a main effect, an interaction with vaccine, an interaction with time point, and the three-way interaction between these variables. Thus, four hypotheses of interest were tested in each model, except where interaction terms were removed to resolve multicollinearity (see Supplement Methods for more detail). Predictor significance was tested using F statistics. Ultimately, 126 p-values (1 to 4 per model) were drawn from 36 models; these were consolidated and corrected for multiple comparisons using the Benjamini-Hochberg method. Statistical significance was defined as corrected p < 0.05; significant F-statistics were followed by post-hoc t-tests without further correction. All presented results represent marginal effects, i.e., effects adjusted for the other terms in the model. Thus, where results are presented without respect to outcome time point, these represent average effects across both time points. For statistically significant continuous predictors, the partial correlation (r_p_) was provided alongside absolute effect sizes. Visualizations represent marginal means (i.e., least-squares means) +/- 95% confidence intervals (CI) along with partial residuals. Detailed information can be found in the Supplement Methods.

## Results

### Sample characteristics

A total of 534 individuals were recruited for the broader study, of whom 364 met criteria for inclusion in the present analyses (Figure 1). Of these, symptom data were collected from 363 subjects and biometric data were collected from 174 subjects. Sample characteristics are provided in Table 1.

**Table 1.**
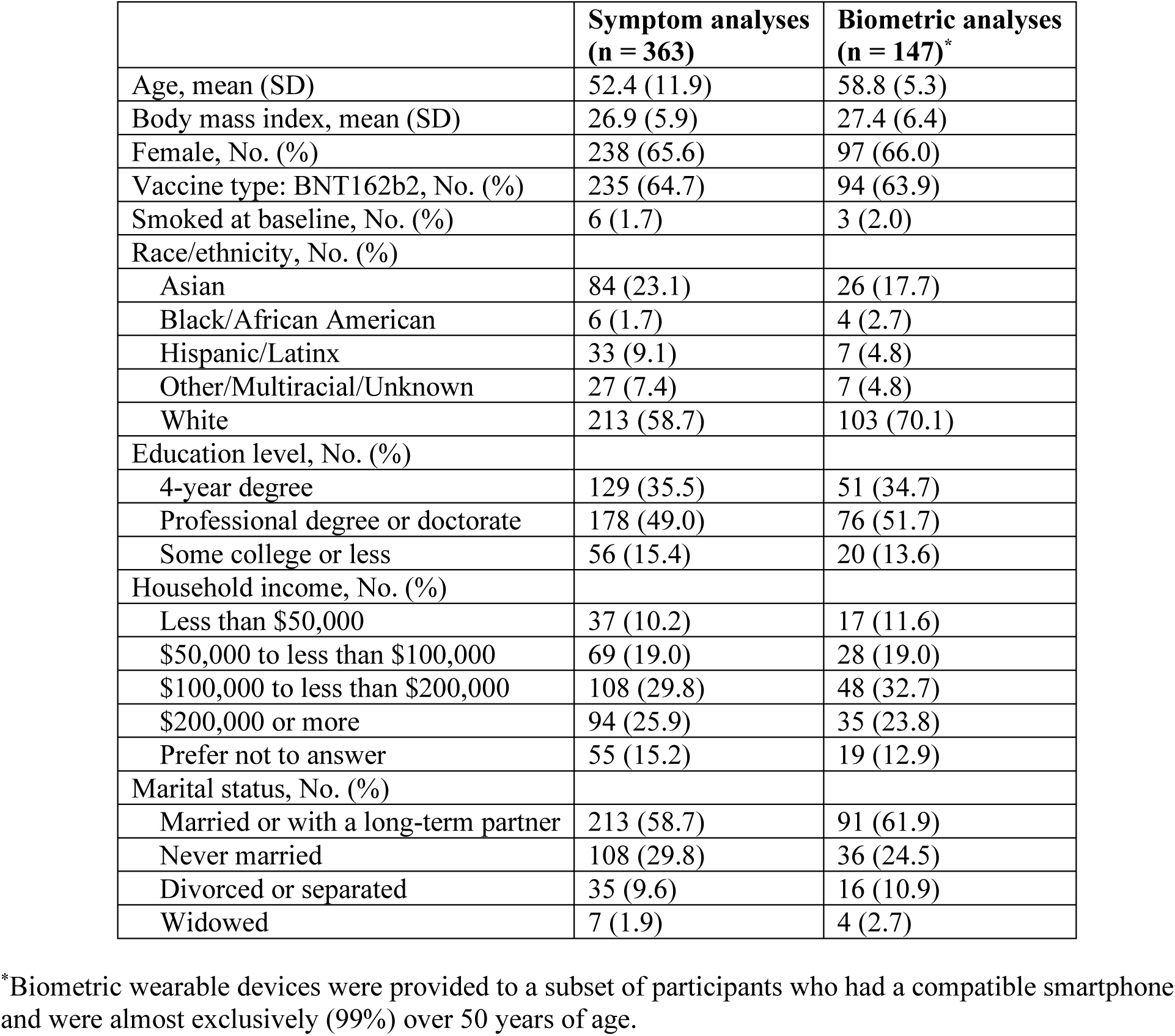
Sample characteristics.

### Symptom predictors of neutralizing antibodies

The frequency of each symptom at each vaccine dose is provided in Supplement Table 3. Test statistics and multiplicity-corrected p-values for all symptom and biometric analyses are provided in Supplement Table 4. Following correction for multiple comparisons, no statistically significant associations were identified between the presence or absence of any symptom at dose one and subsequent nAB. For dose two, main effects were significant for four of 13 symptoms (Figure 2). Specifically, nAB were higher for subjects reporting vs. not reporting the following symptoms at dose two: chills (1.62 fold higher ID50, CI 1.31 to 2.01), feeling unwell (1.48 fold higher ID50, CI 1.22 to 1.79), tiredness (1.47 fold higher ID50, CI 1.17 to 1.83), and headache (1.43 fold higher ID50, CI 1.19 to 1.72). Because symptom presence did not interact with outcome time point or vaccine for any symptom, presented values represent the average association across both vaccines and both outcome time points.

**Figure 2.**
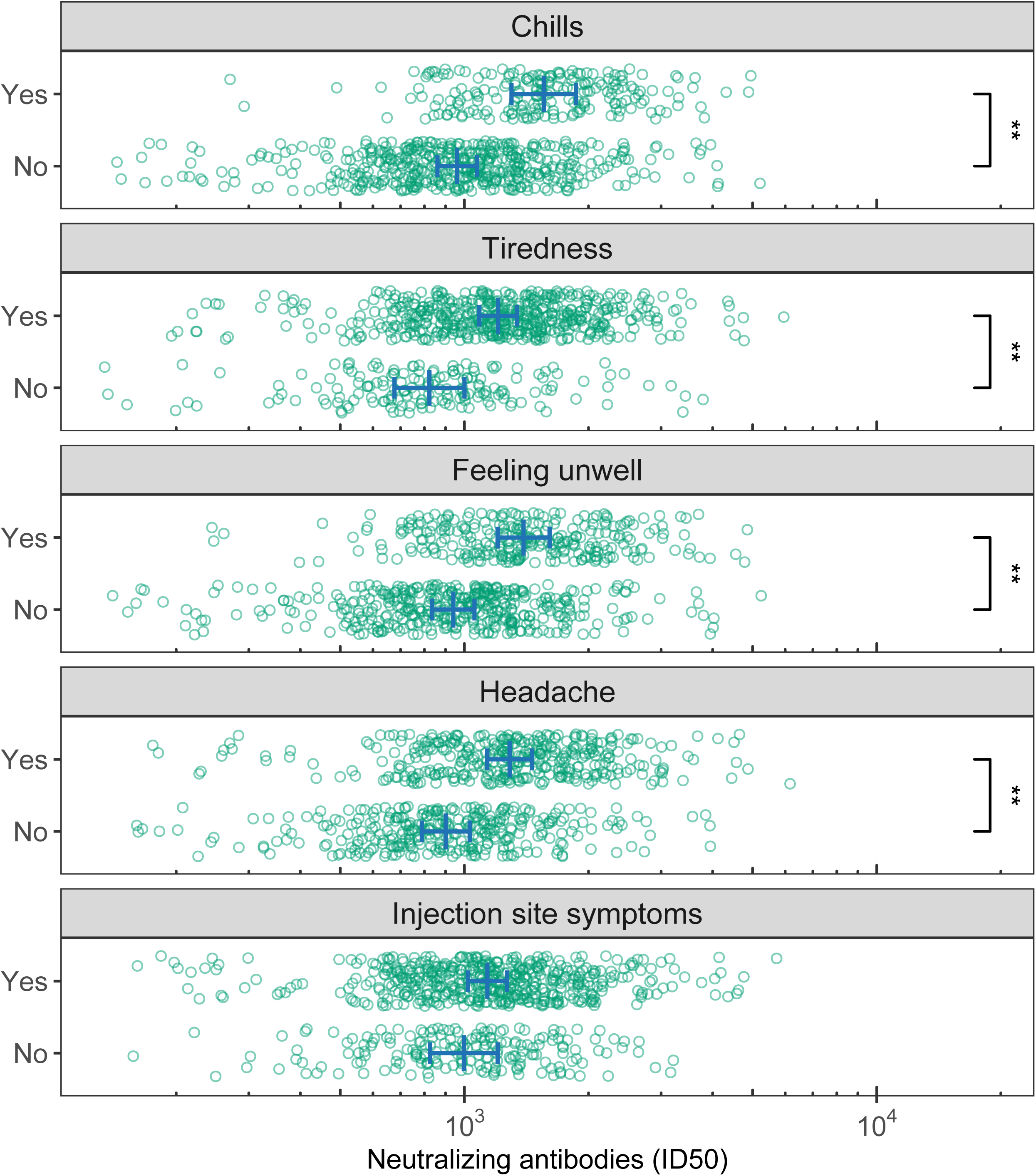
Association between symptoms following the second vaccine dose and subsequent neutralizing antibody levels. Each of 13 symptoms following each vaccine dose was individually tested as a predictor of neutralizing antibody levels at 1 and 6 months later in multivariable mixed-effects models. Neutralizing antibody titer was expressed as the inhibitory dose 50 (ID50). After correcting for multiple comparisons, four symptoms remained statistically significant predictors of neutralizing antibodies, all only when measured at dose two: chills, tiredness, feeling unwell, and headache. Injection site symptoms are included in the figure for comparison. Blue lines represent the marginal means +/- 95% confidence intervals. No interaction terms between a symptom and vaccine or outcome time point were statistically significant; therefore, presented marginal means represent the average effect across both vaccines and both outcome time points. ** = p < 0.01.

### Symptom count as a predictor of neutralizing antibodies

Symptom count was intended as a continuous index of systemic symptom burden, so injection site symptoms were excluded from counting. For dose one, there were no main or interaction effects involving symptom count. For dose two, no interactions were significant, but there was a main effect of symptom count (Figure 3), involving a 1.10 fold higher ID50 (CI 1.06 to 1.14) per additional symptom (partial correlation, r_p_ = 0.27, CI 0.17 to 0.36).

**Figure 3.**
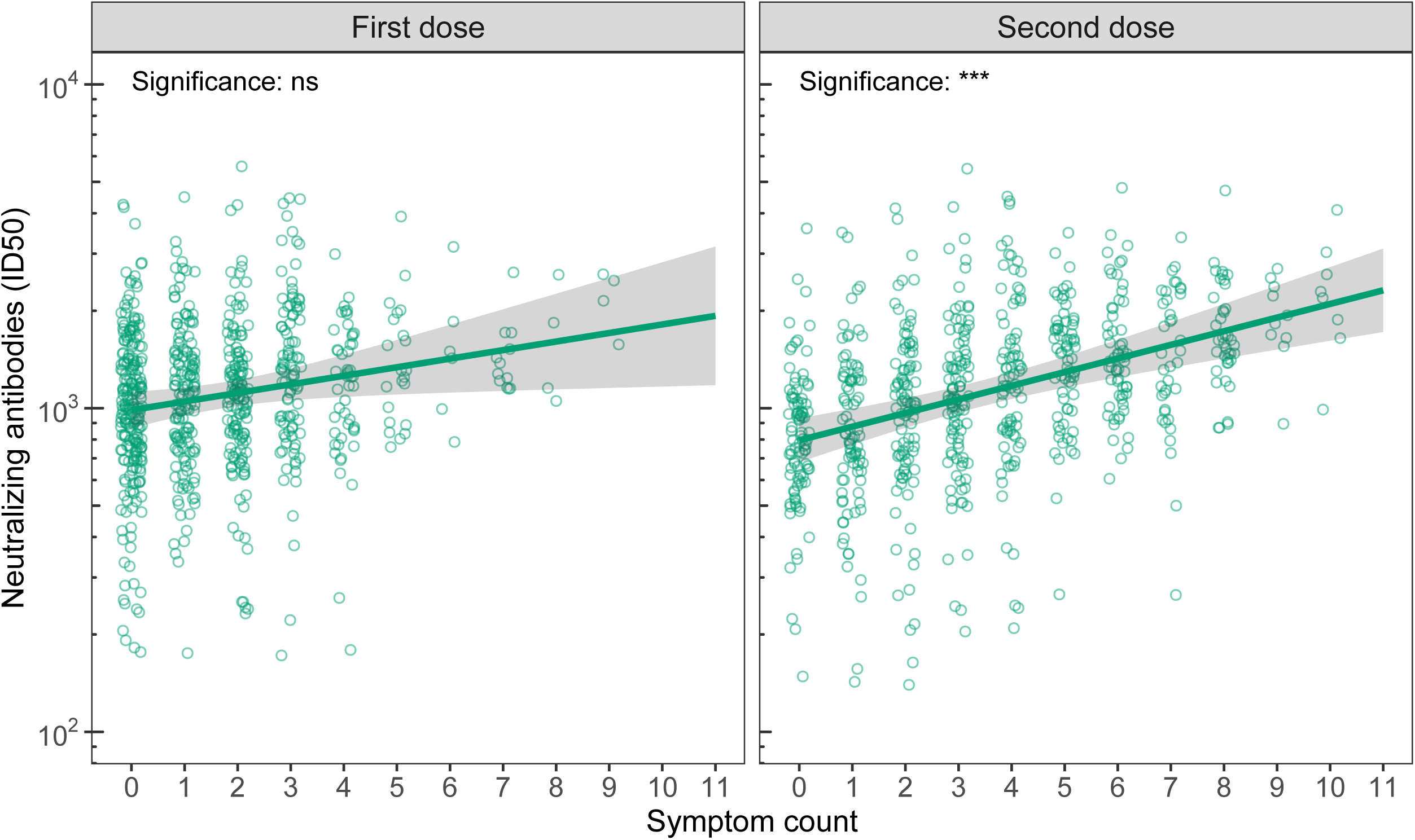
Association between symptom count following each vaccine dose and subsequent neutralizing antibody levels. Symptom count was intended as a measure of systemic symptom burden, so injection site symptoms were excluded from counting. Symptom count following the second dose was a statistically significant predictor of subsequent neutralizing antibody level (p < 0.001). For both doses, there was no significant interaction between symptom count and vaccine or outcome time point (1 month and 6 months following the second dose); therefore, results represent the average relationship across both time points and both vaccines (i.e., marginal means +/- 95% confidence intervals). ID50 = inhibitory dose 50; ns = non-significant; * = p < 0.05; ** = p < 0.01; *** = p < 0.001.

### Biometric predictors of neutralizing antibodies

For vaccination-induced change in nightly 99^th^ percentile skin temperature at dose one, there were no significant main or interaction effects. However, at dose two, there was a significant interaction between outcome time point and vaccination-induced change in nightly 99^th^ percentile skin temperature (Figure 4, top right). Post-hoc testing revealed that vaccination-induced change in skin temperature was predictive of nAB at 1-month follow-up (fold change in ID50 per degree Celsius: 1.84, CI 1.33 to 2.53, p < 0.001; r_p_ = 0.27, CI 0.13 to 0.39) and at 6-month follow-up (fold change in ID50 per degree Celsius: 3.13, CI 2.26 to 4.33, p < 0.001; r_p_ = 0.45, CI 0.33 to 0.55), with the larger effect size at the 6-month follow-up being responsible for the interaction.

**Figure 4.**
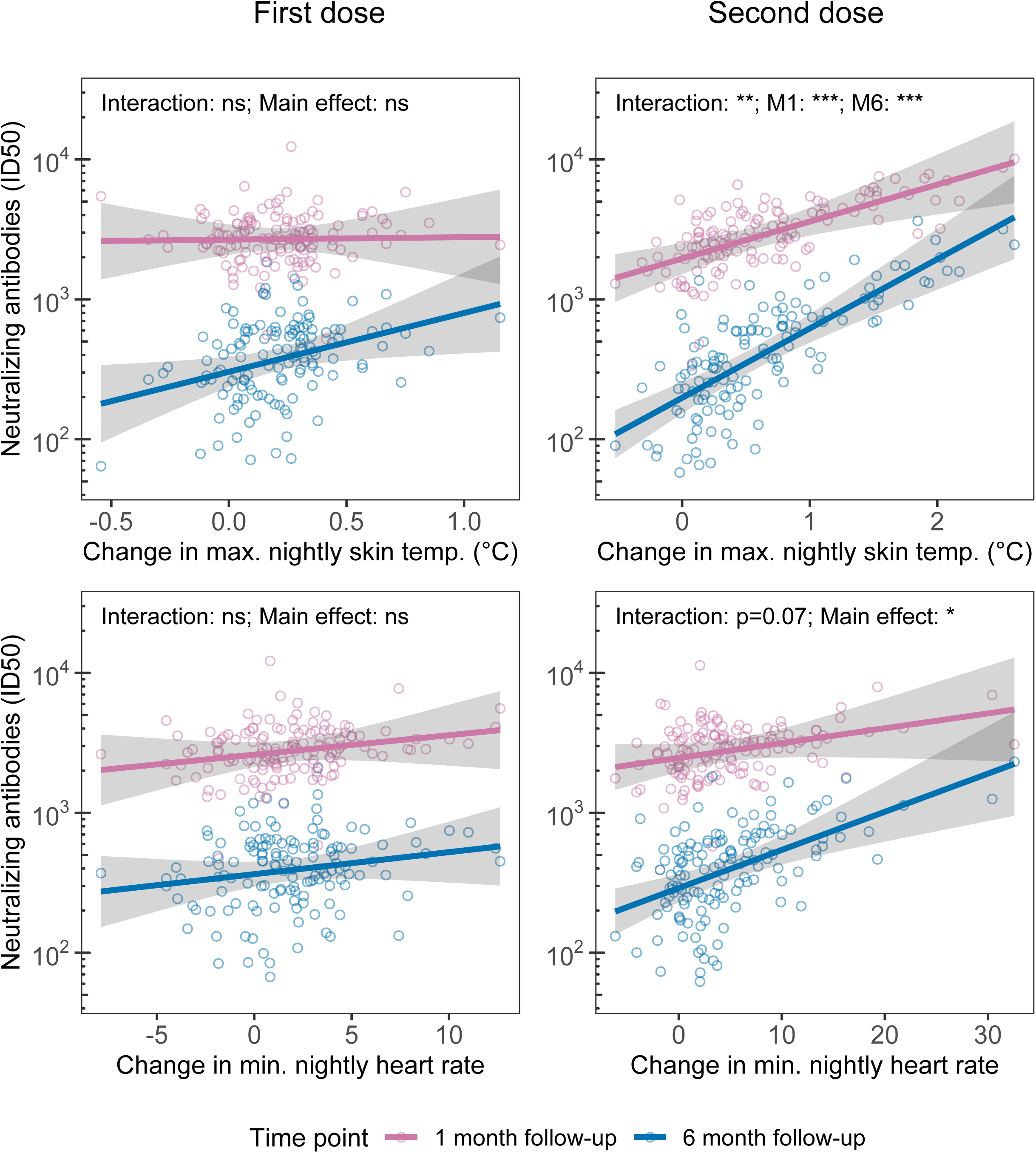
Association between vaccination-induced change in two physiological domains and subsequent neutralizing antibody levels. A subset of study participants wore biometric devices that collected physiological measurements during sleep. Graphs depict the relationship between vaccination-induced change in nightly maximum (99^th^ percentile) skin temperature (top row) and nightly minimum (1^st^ percentile) heart rate (bottom row) and subsequent neutralizing antibody (nAB) level at each outcome time point. Vaccination-induced change in maximum skin temperature at dose two predicted nAB level at both outcome time points, with a stronger association for the 6-month than the 1-month outcome (top right). There was a main effect of vaccination-induced change in minimum heart rate on subsequent nAB level (bottom right). ID50 = inhibitory dose 50; ns = non-significant; * = p < 0.05; ** = p < 0.01; *** = p < 0.001.

For vaccination-induced change in nightly 1^st^ percentile heart rate at dose one, there were no significant main or interaction effects. However, at dose two, a significant main effect of vaccination-induced change in nightly 1^st^ percentile heart rate was observed (Figure 4, bottom right), in the absence of any interaction with outcome time point or vaccine. For each 10 beat per minute increase in heart rate from a subject’s norm, ID50 increased by 1.54 fold (CI 1.18 to 2.02; r_p_ = 0.27, CI 0.10 to 0.41).

Neither vaccination-induced change in nightly 99^th^ percentile heart rate variability nor in average nightly respiratory rate was significantly predictive of subsequent nAB via either main or interaction effects, for either vaccine dose.

## Discussion

We show here that individuals who reported experiencing chills, tiredness, feeling unwell, or headache following the second dose of a SARS-CoV-2 vaccine subsequently had 1.4 to 1.6 times the neutralizing antibody level of people who did not report each symptom, at 1 and 6 months later. We also show that each additional symptom experienced following dose two predicted a 1.1-fold increase in subsequent nAB. This means that, on average, individuals reporting 7 total symptoms subsequently had roughly double the nAB level of individuals reporting 0 symptoms. Using objective biometric data, we present convergent findings showing that greater vaccination-induced change in skin temperature and heart rate, specifically at dose two, predicts greater nAB at both 1 months and 6 months later. Effect sizes were again large, with every 1 degree Celsius of vaccination-induced skin temperature change being associated with a tripling of the nAB level at 6 months later.

Several prior publications have examined the association between systemic symptoms following receipt of a SARS-CoV-2 mRNA vaccine and subsequent nAB level, with inconsistent results. In one report, none of three local or eight systemic symptoms, nor the presence of any local or any systemic symptom, predicted nAB four weeks later (8). By contrast, the presence of at least one systemic symptom was associated with higher nAB at 12-19 days after dose two (10) and at 54 days after dose three (9). There are a few other reports examining the association between reactogenicity to SARS-CoV-2 vaccination and subsequent nAB, but interpretability is limited due to low samples sizes (4 to 8 per condition) (19), the analysis only of nAB trajectories over time (20), which are confounded by absolute initial levels (11), and the use of a mixed sample of mRNA and adenoviral vector vaccine recipients (21).

There are several key strengths of our study compared to prior studies (8–10). Firstly, neither of the previous two studies reporting a significant association between symptoms and nAB examined individual symptoms as predictors. Here, we show that chills, tiredness (or fatigue), feeling unwell, and headache have the strongest predictive relationship with nAB. Secondly, these studies all measured nAB within 2 months of receipt of the second dose of an mRNA vaccine, whereas our report includes measurements as late as 6 months. This long follow-up is important given that after receiving the initial vaccine series, typically a minimum of several months pass before individuals receive a further vaccine dose. Predictive relationships may differ for different outcome time points, and indeed, in the present study, we observed a relationship between vaccination-induced change in skin temperature and nAB that was a stronger predictor of the 6-month than the 1-month outcome. Third, in addition to self-report measures, which might be affected by between-subject differences in the tendency to notice, recall, and report side effects, we use objective biometric measurements of physiological perturbation that are not vulnerable to these influences. Using this data, we present findings that neatly concord with our self-report data. Only one prior study has used non-self-report objective biometric data to predict subsequent humoral immune response after SARS-CoV-2 vaccination (22). That study found positive associations between vaccination-induced change in skin temperature and heart rate and subsequent anti-spike immunoglobulin at roughly 1 month later, in a mixed mRNA and adenoviral vector vaccine sample. Here, we extend those findings, demonstrating similar relationships for nAB and at 6 months, in an mRNA vaccine sample. Finally, our study is among the first to examine associations between symptoms and nAB in a general population sample rather than a convenience sample of healthcare workers.

There are several limitations to our study. Firstly, our results are from individuals who received only the initial COVID-19 vaccine series. It is not clear whether the relationships observed here would apply to individuals undergoing initial vaccination or re-vaccination using updated vaccine formulations. Secondly, our results are from individuals who did not have any serological evidence of SARS-CoV-2 infection. It is unknown whether in individuals with a prior history of SARS-CoV-2 infection, the same predictive ability of symptoms and vaccination-induced change in biometrics would be observed. However, among individuals receiving a two-dose mRNA vaccination, those with a prior history of SARS-CoV-2 infection have been reported to have both greater subsequent anti-spike IgG concentrations (23) and greater reactogenicity (24), suggesting that a predictive association between neutralizing antibodies and reactogenicity may be likely even in previously-infected individuals. A third limitation is that our pseudovirus assay used the spike protein from the original Wuhan/D614G strain of SARS-CoV-2, which may limit generalizability of the findings. Finally, we only address humoral immunity in this study, and although evidence suggests that neutralizing antibodies mediate roughly two-thirds of vaccine efficacy (12), cellular immunity is believed to play an important role in protection from severe disease (25,26).

In sum, we show here in a large community sample that systemic symptoms and increases in skin temperature and heart rate following SARS-CoV-2 mRNA vaccination predict higher subsequent nAB level. We show that these relationships are stronger when predicting long-term rather than short-term nAB outcome. Our findings suggest that a reframing of systemic side effects as desirable may help to address the low rate of ongoing vaccine uptake, given that this appears to be at least partly the result of worry about side effects (4).

## Data Availability

All data produced in the present study are available upon reasonable request to the authors

## Acknowledgements

The project was supported by funding from the National Institutes of Health (R24AG048024, 5U24AG066528, and U54CA260581).

## Author Contributions

ESE, AEM, FMH, JER, SSD, and AAP conceived of and designed the broader study; ESE, AEM, JER, and AAP collected the data; EGD, JER, and AAP accessed and verified data; EGD designed and conducted the statistical analyses; EGD wrote the first draft of the manuscript; ESE, AEM, FMH, JER, SSD, and AAP provided critical feedback on the manuscript.

## Supplement Tables

**Supplement Table 1.**
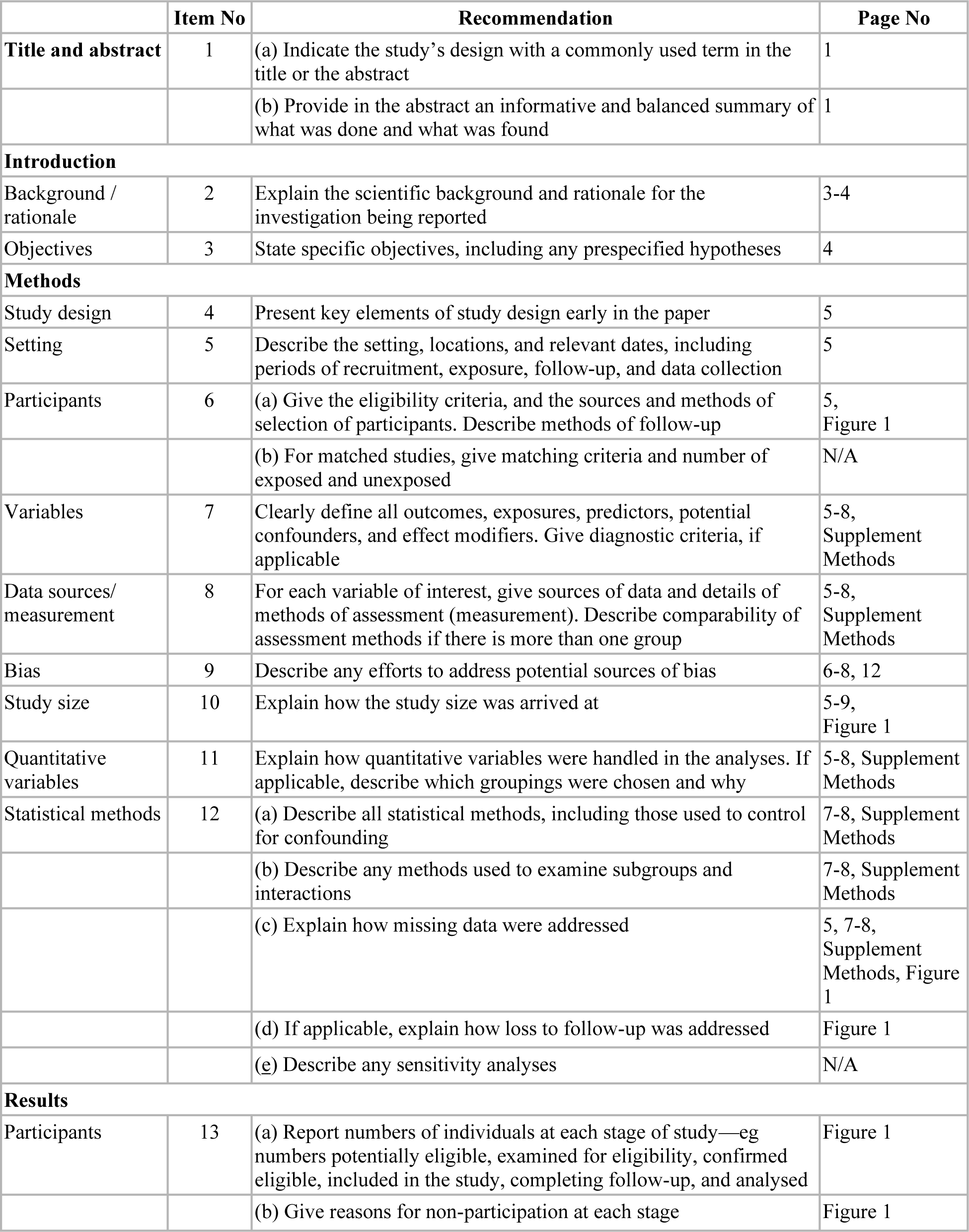
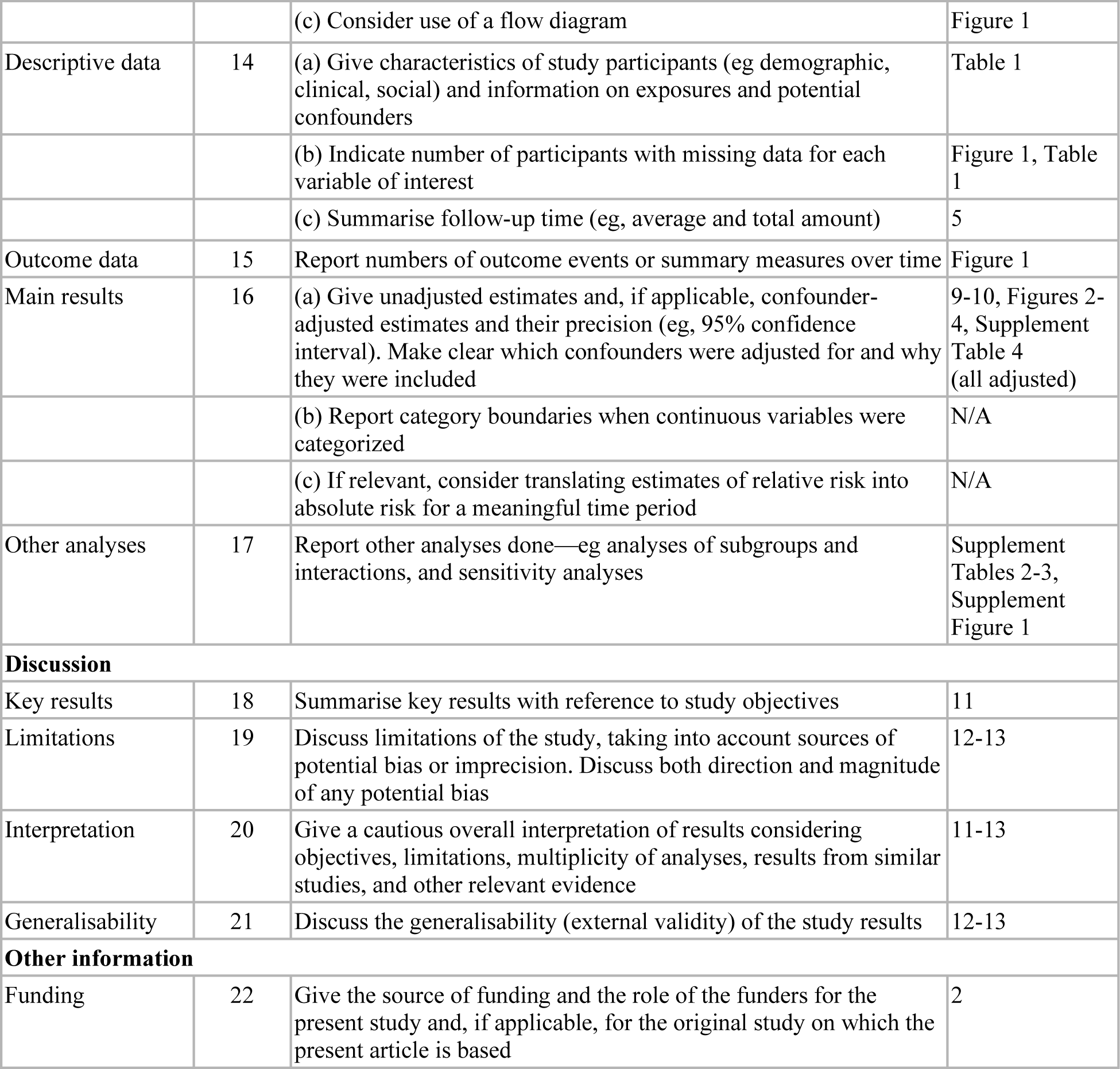
STROBE checklist for cohort studies.

|  | Item No | Recommendation | Page No |
| --- | --- | --- | --- |
| **Title and abstract** | 1 | (a) Indicate the study's design with a commonly used term in the title or the abstract | 1 |
|  |  | (b) Provide in the abstract an informative and balanced summary of what was done and what was found | 1 |
| **Introduction** |  |  |  |
| Background / rationale | 2 | Explain the scientific background and rationale for the investigation being reported | 3-4 |
| Objectives | 3 | State specific objectives, including any prespecified hypotheses | 4 |
| **Methods** |  |  |  |
| Study design | 4 | Present key elements of study design early in the paper | 5 |
| Setting | 5 | Describe the setting, locations, and relevant dates, including periods of recruitment, exposure, follow-up, and data collection | 5 |
| Participants | 6 | (a) Give the eligibility criteria, and the sources and methods of selection of participants. Describe methods of follow-up | 5, Figure 1 |
|  |  | (b) For matched studies, give matching criteria and number of exposed and unexposed | N/A |
| Variables | 7 | Clearly define all outcomes, exposures, predictors, potential confounders, and effect modifiers. Give diagnostic criteria, if applicable | 5-8, Supplement Methods |
| Data sources/ measurement | 8 | For each variable of interest, give sources of data and details of methods of assessment (measurement). Describe comparability of assessment methods if there is more than one group | 5-8, Supplement Methods |
| Bias | 9 | Describe any efforts to address potential sources of bias | 6-8, 12 |
| Study size | 10 | Explain how the study size was arrived at | 5-9, Figure 1 |
| Quantitative variables | 11 | Explain how quantitative variables were handled in the analyses. If applicable, describe which groupings were chosen and why | 5-8, Supplement Methods |
| Statistical methods | 12 | (a) Describe all statistical methods, including those used to control for confounding | 7-8, Supplement Methods |
|  |  | (b) Describe any methods used to examine subgroups and interactions | 7-8, Supplement Methods |
|  |  | (c) Explain how missing data were addressed | 5, 7-8, Supplement Methods, Figure 1 |
|  |  | (d) If applicable, explain how loss to follow-up was addressed | Figure 1 |
|  |  | (e) Describe any sensitivity analyses | N/A |
| **Results** |  |  |  |
| Participants | 13 | (a) Report numbers of individuals at each stage of study—eg numbers potentially eligible, examined for eligibility, confirmed eligible, included in the study, completing follow-up, and analysed | Figure 1 |
|  |  | (b) Give reasons for non-participation at each stage | Figure 1 |
|  |  | (c) Consider use of a flow diagram | Figure 1 |
| Descriptive data | 14 | (a) Give characteristics of study participants (eg demographic, clinical, social) and information on exposures and potential confounders | Table 1 |
|  |  | (b) Indicate number of participants with missing data for each variable of interest | Figure 1, Table 1 |
|  |  | (c) Summarise follow-up time (eg, average and total amount) | 5 |
| Outcome data | 15 | Report numbers of outcome events or summary measures over time | Figure 1 |
| Main results | 16 | (a) Give unadjusted estimates and, if applicable, confounder-adjusted estimates and their precision (eg, 95% confidence interval). Make clear which confounders were adjusted for and why they were included | 9-10, Figures 2-4, Supplement Table 4 (all adjusted) |
|  |  | (b) Report category boundaries when continuous variables were categorized | N/A |
|  |  | (c) If relevant, consider translating estimates of relative risk into absolute risk for a meaningful time period | N/A |
| Other analyses | 17 | Report other analyses done—eg analyses of subgroups and interactions, and sensitivity analyses | Supplement Tables 2-3, Supplement Figure 1 |
| **Discussion** |  |  |  |
| Key results | 18 | Summarise key results with reference to study objectives | 11 |
| Limitations | 19 | Discuss limitations of the study, taking into account sources of potential bias or imprecision. Discuss both direction and magnitude of any potential bias | 12-13 |
| Interpretation | 20 | Give a cautious overall interpretation of results considering objectives, limitations, multiplicity of analyses, results from similar studies, and other relevant evidence | 11-13 |
| Generalisability | 21 | Discuss the generalisability (external validity) of the study results | 12-13 |
| **Other information** |  |  |  |
| Funding | 22 | Give the source of funding and the role of the funders for the present study and, if applicable, for the original study on which the present article is based | 2 |

**Supplement Table 2.** Descriptive statistics for biometric data.

| **Vaccination-induced change in:** | **Dose one mean (SD); n = 141** | **Dose two mean (SD); n = 141** | **One-sample t-statistic for dose two** | **p value** |
| --- | --- | --- | --- | --- |
| HR: nightly 99th percentile | 2.81 (4.95) | 6.19 (7.72) | 9.51 | 7e-17 |
| HR: nightly mean | 2.12 (3.74) | 5.48 (6.89) | 9.45 | 1e-16 |
| HR: nightly 1st percentile | 1.82 (3.27) | 4.76 (6.04) | 9.36 | 2e-16 |
| HRV: nightly 99th percentile | -9.05 (13.62) | -10.63 (15.19) | -8.30 | 8e-14 |
| HRV: nightly mean | -4.44 (8.12) | -5.6 (8.62) | -7.72 | 2e-12 |
| HRV: nightly 1st percentile | -2.74 (5.74) | -3.11 (5.96) | -6.20 | 6e-09 |
| ST: nightly 99th percentile | 0.21 (0.23) | 0.63 (0.63) | 11.67 | 5e-22 |
| ST: nightly mean | 0.27 (0.39) | 0.48 (0.5) | 11.09 | 1e-20 |
| ST: nightly 1st percentile | 0.71 (1.92) | 0.76 (1.88) | 4.69 | 7e-06 |
| RR: nightly mean | 0.3 (0.56) | 0.94 (1.01) | 11.00 | 1e-20 |

Biometric data at the time of either vaccine dose was available for a total of 147 subjects, with 141 providing data at dose one and 141 providing data at dose two. Respiratory rate (RR; breaths per minute) was available only as a nightly average. For heart rate (HR; beats per minute), heart rate variability (HRV; root mean square of successive differences, in milliseconds), and skin temperature (ST; degrees Celsius), one-sample t-tests were used to identify the approach to summarizing the nightly time series that was most sensitive to vaccination-induced change. For HRV and ST, this was the 99<sup>th</sup> percentile, and for HR, it was the 1<sup>st</sup> percentile. These summary approaches were then used to test hypotheses regarding the ability of vaccination-induced change in each physiological domain to predict subsequent neutralizing antibody level.

**Supplement Table 3.**
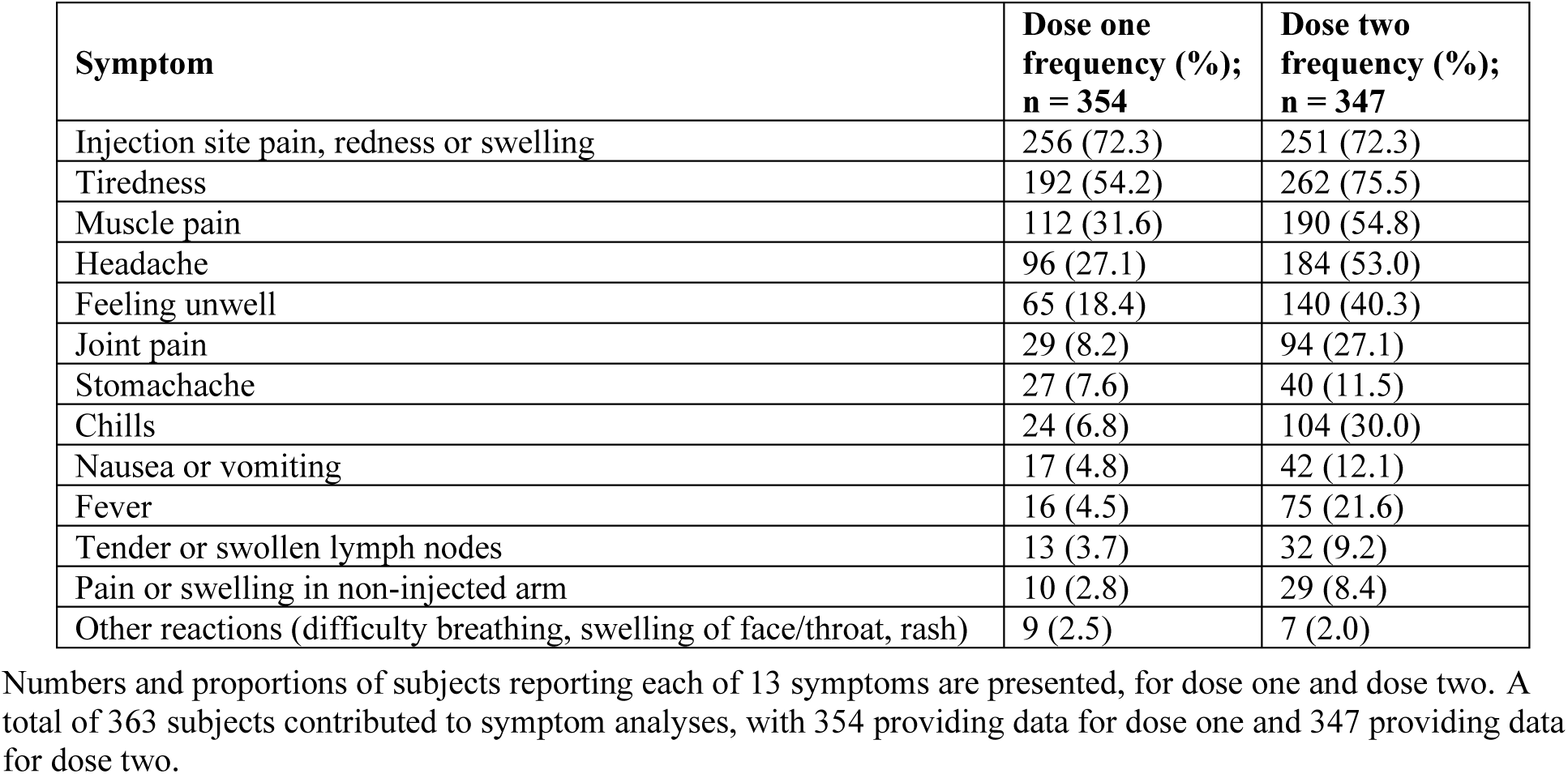
Descriptive statistics for symptoms.

| **Symptom** | **Dose one**<br>**frequency (%);**<br>**n = 354** | **Dose two**<br>**frequency (%);**<br>**n = 347** |
| --- | --- | --- |
| Injection site pain, redness or swelling | 256 (72.3) | 251 (72.3) |
| Tiredness | 192 (54.2) | 262 (75.5) |
| Muscle pain | 112 (31.6) | 190 (54.8) |
| Headache | 96 (27.1) | 184 (53.0) |
| Feeling unwell | 65 (18.4) | 140 (40.3) |
| Joint pain | 29 (8.2) | 94 (27.1) |
| Stomachache | 27 (7.6) | 40 (11.5) |
| Chills | 24 (6.8) | 104 (30.0) |
| Nausea or vomiting | 17 (4.8) | 42 (12.1) |
| Fever | 16 (4.5) | 75 (21.6) |
| Tender or swollen lymph nodes | 13 (3.7) | 32 (9.2) |
| Pain or swelling in non-injected arm | 10 (2.8) | 29 (8.4) |
| Other reactions (difficulty breathing, swelling of face/throat, rash) | 9 (2.5) | 7 (2.0) |

Numbers and proportions of subjects reporting each of 13 symptoms are presented, for dose one and dose two. A total of 363 subjects contributed to symptom analyses, with 354 providing data for dose one and 347 providing data for dose two.

**Supplement Table 4.** Test statistics and effect sizes for all examined predictor variables.

| Predictor | Partial $\eta^2$ | F | Df | p<br>(uncorr.) | p | |
| --- | --- | --- | --- | --- | --- | --- |
| Change in max. nightly skin temp. (D2) | 0.20 (0.09 to 0.32) | 31.30 | 1, 125.1 | 1e-07 | 2e-05 | *** |
| Symptom count (D2) | 0.07 (0.03 to 0.12) | 24.50 | 1, 337.9 | 1e-06 | 7e-05 | *** |
| Chills (D2) | 0.05 (0.01 to 0.10) | 17.78 | 1, 338.0 | 3e-05 | 0.001 | ** |
| Feeling unwell (D2) | 0.04 (0.01 to 0.09) | 15.75 | 1, 338.7 | 9e-05 | 0.003 | ** |
| Headache (D2) | 0.04 (0.01 to 0.09) | 14.56 | 1, 338.9 | 2e-04 | 0.004 | ** |
| Tiredness (D2) | 0.04 (0.01 to 0.09) | 14.38 | 1, 339.3 | 2e-04 | 0.004 | ** |
| Time point × Change in max. nightly skin temp. (D2) | 0.10 (0.02 to 0.21) | 13.96 | 1, 127.5 | 3e-04 | 0.005 | ** |
| Change in min. nightly HR (D2) | 0.07 (0.01 to 0.17) | 10.20 | 1, 132.0 | 0.002 | 0.028 | * |
| Time point × Change in avg. nightly RR (D1) | 0.06 (0.01 to 0.15) | 8.60 | 1, 134.4 | 0.004 | 0.055 | |
| Time point × Change in avg. nightly RR (D2) | 0.06 (0.01 to 0.15) | 8.08 | 1, 134.6 | 0.005 | 0.065 | |
| Tiredness (D1) | 0.02 (0.00 to 0.06) | 7.68 | 1, 345.9 | 0.006 | 0.067 |  |
| Time point × Change in min. nightly HR (D2) | 0.05 (0.00 to 0.14) | 7.60 | 1, 134.2 | 0.007 | 0.070 | |
| Vaccine × Time point × Change in min. nightly HR (D1) | 0.05 (0.00 to 0.14) | 7.16 | 1, 134.4 | 0.008 | 0.081 | |
| Change in avg. nightly RR (D2) | 0.05 (0.00 to 0.13) | 6.51 | 1, 132.2 | 0.012 | 0.10 |  |
| Time point × Change in max. nightly skin temp. (D1) | 0.05 (0.00 to 0.14) | 6.40 | 1, 127.2 | 0.013 | 0.10 | |
| Tender or swollen lymph nodes (D2) | 0.02 (0.00 to 0.05) | 6.22 | 1, 342.5 | 0.013 | 0.10 |  |
| Joint pain (D2) | 0.02 (0.00 to 0.05) | 5.85 | 1, 338.2 | 0.016 | 0.12 |  |
| Muscle pain (D2) | 0.02 (0.00 to 0.05) | 5.63 | 1, 338.2 | 0.018 | 0.13 |  |
| Fever (D2) | 0.02 (0.00 to 0.05) | 5.21 | 1, 336.9 | 0.023 | 0.15 |  |
| Symptom count (D1) | 0.01 (0.00 to 0.05) | 4.81 | 1, 344.7 | 0.029 | 0.18 |  |
| Vaccine × Change in max. nightly skin temp. (D2) | 0.04 (0.00 to 0.12) | 4.70 | 1, 125.3 | 0.032 | 0.19 | |
| Vaccine × Symptom count (D2) | 0.01 (0.00 to 0.04) | 3.99 | 1, 337.7 | 0.047 | 0.27 | |
| Vaccine × Feeling unwell (D2) | 0.01 (0.00 to 0.04) | 3.76 | 1, 339.7 | 0.053 | 0.29 | |
| Vaccine × Tender or swollen lymph nodes (D2) | 0.01 (0.00 to 0.04) | 3.46 | 1, 343.7 | 0.064 | 0.33 | |
| Tender or swollen lymph nodes (D1) | 0.01 (0.00 to 0.04) | 3.43 | 1, 344.7 | 0.065 | 0.33 |  |
| Vaccine × Joint pain (D2) | 0.01 (0.00 to 0.04) | 3.26 | 1, 338.2 | 0.072 | 0.35 | |
| Other allergic reactions (difficulty breathing, swelling of face/throat, rash) (D1) | 0.01 (0.00 to 0.04) | 3.14 | 1, 344.8 | 0.077 | 0.36 |  |
| Time point × Joint pain (D2) | 0.01 (0.00 to 0.04) | 3.02 | 1, 337.5 | 0.083 | 0.37 | |
| Vaccine × Time point × Pain or swelling in non-injected arm (D2) | 0.01 (0.00 to 0.04) | 2.88 | 1, 343.1 | 0.091 | 0.39 | |
| Vaccine × Feeling unwell (D1) | 0.01 (0.00 to 0.04) | 2.85 | 1, 345.8 | 0.092 | 0.39 | |
| Vaccine × Chills (D2) | 0.01 (0.00 to 0.04) | 2.63 | 1, 338.1 | 0.11 | 0.43 | |
| Vaccine × Time point × Muscle pain (D1) | 0.01 (0.00 to 0.04) | 2.59 | 1, 345.2 | 0.11 | 0.43 | |
| Vaccine × Joint pain (D1) | 0.01 (0.00 to 0.03) | 2.51 | 1, 354.8 | 0.11 | 0.43 | |
| Vaccine × Pain or swelling in non-injected arm (D2) | 0.01 (0.00 to 0.03) | 2.32 | 1, 343.7 | 0.13 | 0.48 | |
| Vaccine × Fever (D2) | 0.01 (0.00 to 0.03) | 2.14 | 1, 337.1 | 0.14 | 0.51 | |
| Vaccine × Change in max. nightly HRV (D1) | 0.02 (0.00 to 0.08) | 2.15 | 1, 132.0 | 0.15 | 0.51 | |
| Injection site pain, redness or swelling (D2) | 0.01 (0.00 to 0.03) | 2.03 | 1, 337.5 | 0.16 | 0.53 |  |
| Time point × Muscle pain (D1) | 0.01 (0.00 to 0.03) | 1.87 | 1, 345.2 | 0.17 | 0.53 | |
| Vaccine × Time point × Injection site pain, redness or swelling (D2) | 0.01 (0.00 to 0.03) | 1.83 | 1, 336.4 | 0.18 | 0.53 | |
| Vaccine × Time point × Stomachache (D1) | 0.01 (0.00 to 0.03) | 1.81 | 1, 344.4 | 0.18 | 0.53 | |
| Vaccine × Change in min. nightly HR (D1) | 0.01 (0.00 to 0.08) | 1.76 | 1, 132.3 | 0.19 | 0.53 | |
| Vaccine × Time point × Change in max. nightly HRV (D1) | 0.01 (0.00 to 0.07) | 1.74 | 1, 134.1 | 0.19 | 0.53 | |
| Time point × Feeling unwell (D2) | 0.01 (0.00 to 0.03) | 1.72 | 1, 337.8 | 0.19 | 0.53 | |
| Vaccine × Time point × Joint pain (D2) | 0.01 (0.00 to 0.03) | 1.70 | 1, 337.9 | 0.19 | 0.53 | |
| Change in min. nightly HR (D1) | 0.01 (0.00 to 0.07) | 1.69 | 1, 132.4 | 0.20 | 0.53 |  |
| Headache (D1) | 0.00 (0.00 to 0.03) | 1.68 | 1, 345.3 | 0.20 | 0.53 |  |
| Time point × Joint pain (D1) | 0.00 (0.00 to 0.03) | 1.68 | 1, 351.2 | 0.20 | 0.53 | |
| Time point × Stomachache (D2) | 0.00 (0.00 to 0.03) | 1.61 | 1, 335.8 | 0.21 | 0.54 | |
| Change in max. nightly skin temp. (D1) | 0.01 (0.00 to 0.08) | 1.53 | 1, 125.2 | 0.22 | 0.56 |  |
| Muscle pain (D1) | 0.00 (0.00 to 0.03) | 1.39 | 1, 344.5 | 0.24 | 0.59 |  |
| Vaccine × Chills (D1) | 0.00 (0.00 to 0.03) | 1.37 | 1, 343.9 | 0.24 | 0.59 | |
| Vaccine × Tiredness (D2) | 0.00 (0.00 to 0.03) | 1.35 | 1, 342.8 | 0.25 | 0.59 | |
| Vaccine × Symptom count (D1) | 0.00 (0.00 to 0.03) | 1.34 | 1, 344.5 | 0.25 | 0.59 | |
| Other allergic reactions (difficulty breathing, swelling of face/throat, rash) (D2) | 0.00 (0.00 to 0.03) | 1.30 | 1, 369.8 | 0.25 | 0.59 |  |
| Vaccine × Time point × Change in max. nightly HRV (D2) | 0.01 (0.00 to 0.07) | 1.30 | 1, 134.1 | 0.26 | 0.59 | |
| Vaccine × Change in max. nightly skin temp. (D1) | 0.01 (0.00 to 0.07) | 1.11 | 1, 124.4 | 0.29 | 0.66 | |
| Vaccine × Time point × Muscle pain (D2) | 0.00 (0.00 to 0.03) | 1.07 | 1, 337.7 | 0.30 | 0.67 | |
| Change in avg. nightly RR (D1) | 0.01 (0.00 to 0.06) | 1.01 | 1, 132.3 | 0.32 | 0.69 |  |
| Time point × Symptom count (D2) | 0.00 (0.00 to 0.02) | 0.92 | 1, 337.1 | 0.34 | 0.70 | |
| Time point × Muscle pain (D2) | 0.00 (0.00 to 0.02) | 0.90 | 1, 337.3 | 0.34 | 0.70 | |
| Nausea vomiting (D1) | 0.00 (0.00 to 0.02) | 0.90 | 1, 345.1 | 0.34 | 0.70 |  |
| Time point × Chills (D2) | 0.00 (0.00 to 0.02) | 0.88 | 1, 337.8 | 0.35 | 0.70 | |
| Time point × Headache (D2) | 0.00 (0.00 to 0.02) | 0.87 | 1, 338.5 | 0.35 | 0.70 | |
| Vaccine × Time point × Headache (D1) | 0.00 (0.00 to 0.02) | 0.82 | 1, 345.3 | 0.37 | 0.71 | |
| Vaccine × Headache (D2) | 0.00 (0.00 to 0.02) | 0.80 | 1, 339.4 | 0.37 | 0.71 | |
| Chills (D1) | 0.00 (0.00 to 0.02) | 0.80 | 1, 343.8 | 0.37 | 0.71 |  |
| Time point × Stomachache (D1) | 0.00 (0.00 to 0.02) | 0.78 | 1, 344.4 | 0.38 | 0.71 | |
| Vaccine × Stomachache (D2) | 0.00 (0.00 to 0.02) | 0.76 | 1, 336.4 | 0.38 | 0.71 | |
| Time point × Tiredness (D1) | 0.00 (0.00 to 0.02) | 0.68 | 1, 346.7 | 0.41 | 0.75 | |
| Vaccine × Injection site pain, redness or swelling (D2) | 0.00 (0.00 to 0.02) | 0.66 | 1, 337.0 | 0.42 | 0.75 | |
| Vaccine × Tiredness (D1) | 0.00 (0.00 to 0.02) | 0.59 | 1, 346.2 | 0.44 | 0.79 | |
| Vaccine × Muscle pain (D1) | 0.00 (0.00 to 0.02) | 0.54 | 1, 344.8 | 0.46 | 0.80 | |
| Vaccine × Time point × Nausea vomiting (D2) | 0.00 (0.00 to 0.02) | 0.53 | 1, 336.0 | 0.47 | 0.80 | |
| Vaccine × Injection site pain, redness or swelling (D1) | 0.00 (0.00 to 0.02) | 0.52 | 1, 344.4 | 0.47 | 0.80 | |
| Time point × Symptom count (D1) | 0.00 (0.00 to 0.02) | 0.44 | 1, 345.4 | 0.51 | 0.84 | |
| Time point × Pain or swelling in non-injected arm (D2) | 0.00 (0.00 to 0.02) | 0.43 | 1, 339.2 | 0.51 | 0.84 | |
| Vaccine × Time point × Symptom count (D1) | 0.00 (0.00 to 0.02) | 0.42 | 1, 345.2 | 0.52 | 0.84 | |
| Pain or swelling in non-injected arm (D2) | 0.00 (0.00 to 0.02) | 0.41 | 1, 339.6 | 0.52 | 0.84 |  |
| Change in max. nightly HRV (D1) | 0.00 (0.00 to 0.05) | 0.40 | 1, 132.0 | 0.53 | 0.84 |  |
| Stomachache (D2) | 0.00 (0.00 to 0.02) | 0.37 | 1, 336.8 | 0.54 | 0.85 |  |
| Vaccine × Time point × Change in max. nightly skin temp. (D1) | 0.00 (0.00 to 0.05) | 0.29 | 1, 126.6 | 0.59 | 0.89 | |
| Vaccine × Change in avg. nightly RR (D1) | 0.00 (0.00 to 0.04) | 0.29 | 1, 132.2 | 0.59 | 0.89 | |
| Nausea vomiting (D2) | 0.00 (0.00 to 0.02) | 0.27 | 1, 336.4 | 0.61 | 0.89 |  |
| Vaccine × Stomachache (D1) | 0.00 (0.00 to 0.02) | 0.26 | 1, 343.8 | 0.61 | 0.89 | |
| Vaccine × Time point × Tiredness (D1) | 0.00 (0.00 to 0.02) | 0.25 | 1, 347.0 | 0.61 | 0.89 | |
| Time point × Headache (D1) | 0.00 (0.00 to 0.02) | 0.25 | 1, 345.7 | 0.62 | 0.89 | |
| Vaccine × Time point × Chills (D1) | 0.00 (0.00 to 0.02) | 0.24 | 1, 344.5 | 0.62 | 0.89 | |
| Vaccine × Time point × Injection site pain, redness or swelling (D1) | 0.00 (0.00 to 0.02) | 0.24 | 1, 345.0 | 0.62 | 0.89 | |
| Vaccine × Time point × Change in avg. nightly RR (D1) | 0.00 (0.00 to 0.04) | 0.23 | 1, 134.3 | 0.63 | 0.89 | |
| Time point × Change in max. nightly HRV (D1) | 0.00 (0.00 to 0.04) | 0.22 | 1, 134.2 | 0.64 | 0.89 | |
| Vaccine × Time point × Headache (D2) | 0.00 (0.00 to 0.02) | 0.21 | 1, 340.1 | 0.64 | 0.89 | |
| Fever (D1) | 0.00 (0.00 to 0.02) | 0.20 | 1, 344.7 | 0.65 | 0.89 |  |
| Joint pain (D1) | 0.00 (0.00 to 0.02) | 0.20 | 1, 350.5 | 0.65 | 0.89 |  |
| Time point × Injection site pain, redness or swelling (D1) | 0.00 (0.00 to 0.02) | 0.17 | 1, 345.0 | 0.68 | 0.90 | |
| Vaccine × Nausea vomiting (D2) | 0.00 (0.00 to 0.02) | 0.16 | 1, 336.7 | 0.69 | 0.90 | |
| Vaccine × Time point × Tender or swollen lymph nodes (D2) | 0.00 (0.00 to 0.01) | 0.15 | 1, 343.6 | 0.70 | 0.90 | |
| Vaccine × Time point × Change in min. nightly HR (D2) | 0.00 (0.00 to 0.04) | 0.15 | 1, 134.2 | 0.70 | 0.90 | |
| Vaccine × Time point × Joint pain (D1) | 0.00 (0.00 to 0.01) | 0.15 | 1, 355.8 | 0.70 | 0.90 | |
| Pain or swelling in non-injected arm (D1) | 0.00 (0.00 to 0.01) | 0.14 | 1, 344.7 | 0.71 | 0.90 |  |
| Vaccine × Change in min. nightly HR (D2) | 0.00 (0.00 to 0.04) | 0.14 | 1, 132.0 | 0.71 | 0.90 | |
| Time point × Tiredness (D2) | 0.00 (0.00 to 0.02) | 0.12 | 1, 338.2 | 0.73 | 0.91 | |
| Vaccine × Change in avg. nightly RR (D2) | 0.00 (0.00 to 0.03) | 0.10 | 1, 132.3 | 0.76 | 0.93 | |
| Vaccine × Headache (D1) | 0.00 (0.00 to 0.01) | 0.09 | 1, 344.6 | 0.77 | 0.93 | |
| Time point × Feeling unwell (D1) | 0.00 (0.00 to 0.01) | 0.08 | 1, 346.1 | 0.77 | 0.93 | |
| Time point × Change in min. nightly HR (D1) | 0.00 (0.00 to 0.03) | 0.07 | 1, 134.5 | 0.79 | 0.93 | |
| Change in max. nightly HRV (D2) | 0.00 (0.00 to 0.03) | 0.06 | 1, 132.0 | 0.80 | 0.93 |  |
| Time point × Injection site pain, redness or swelling (D2) | 0.00 (0.00 to 0.01) | 0.06 | 1, 336.8 | 0.80 | 0.93 | |
| Vaccine × Time point × Tiredness (D2) | 0.00 (0.00 to 0.01) | 0.06 | 1, 341.1 | 0.80 | 0.93 | |
| Vaccine × Time point × Symptom count (D2) | 0.00 (0.00 to 0.01) | 0.06 | 1, 337.4 | 0.81 | 0.93 | |
| Vaccine × Time point × Feeling unwell (D1) | 0.00 (0.00 to 0.01) | 0.05 | 1, 347.8 | 0.82 | 0.93 | |
| Vaccine × Change in max. nightly HRV (D2) | 0.00 (0.00 to 0.03) | 0.05 | 1, 132.0 | 0.82 | 0.93 | |
| Time point × Nausea vomiting (D2) | 0.00 (0.00 to 0.01) | 0.05 | 1, 335.8 | 0.83 | 0.93 | |
| Feeling unwell (D1) | 0.00 (0.00 to 0.01) | 0.05 | 1, 344.9 | 0.83 | 0.93 |  |
| Vaccine × Time point × Chills (D2) | 0.00 (0.00 to 0.01) | 0.04 | 1, 338.4 | 0.84 | 0.93 | |
| Vaccine × Time point × Stomachache (D2) | 0.00 (0.00 to 0.01) | 0.04 | 1, 335.9 | 0.85 | 0.93 | |
| Vaccine × Time point × Change in avg. nightly RR (D2) | 0.00 (0.00 to 0.03) | 0.03 | 1, 134.5 | 0.86 | 0.93 | |
| Stomachache (D1) | 0.00 (0.00 to 0.01) | 0.03 | 1, 343.8 | 0.87 | 0.93 |  |
| Vaccine × Time point × Feeling unwell (D2) | 0.00 (0.00 to 0.01) | 0.02 | 1, 338.9 | 0.89 | 0.93 | |
| Time point × Tender or swollen lymph nodes (D2) | 0.00 (0.00 to 0.01) | 0.02 | 1, 341.6 | 0.89 | 0.93 | |
| Time point × Chills (D1) | 0.00 (0.00 to 0.01) | 0.02 | 1, 344.5 | 0.89 | 0.93 | |
| Vaccine × Time point × Change in max. nightly skin temp. (D2) | 0.00 (0.00 to 0.02) | 0.02 | 1, 127.5 | 0.90 | 0.93 | |
| Time point × Fever (D2) | 0.00 (0.00 to 0.01) | 0.01 | 1, 336.7 | 0.90 | 0.93 | |
| Vaccine × Time point × Fever (D2) | 0.00 (0.00 to 0.01) | 0.01 | 1, 336.9 | 0.91 | 0.93 | |
| Injection site pain, redness or swelling (D1) | 0.00 (0.00 to 0.01) | 0.01 | 1, 344.4 | 0.92 | 0.94 |  |
| Vaccine × Muscle pain (D2) | 0.00 (0.00 to 0.00) | 0.00 | 1, 338.3 | 0.98 | 0.99 | |
| Time point × Change in max. nightly HRV (D2) | 0.00 (0.00 to 0.00) | 0.00 | 1, 134.2 | 0.99 | 0.99 | |

18 variables (13 symptoms or symptom categories, 1 symptom count, and 4 biometric variables) measured following receipt of vaccine doses one and two were examined as potential predictors of subsequent neutralizing antibodies measured at 1 month and 6 months following dose two, via 36 individual mixed-effects linear models. Where possible, these models included interaction terms involving the variable under investigation with vaccine, outcome time point, and the three-way interaction between these variables. Statistics were extracted from these models and then all 126 p-values were simultaneously corrected for multiple comparisons using the Benjamini-Hochberg method. \* = $p < 0.05$ ; \*\* = $p < 0.01$ ; \*\*\* = $p < 0.001$ .

## Supplement Methods

### Biometric data processing

Collected data were exported from the web application Oura Teams in March 2022. Heart rate (HR) and heart rate variability (HRV) were available as time series of observations in 5-minute intervals. HRV was provided in the form of the root mean square of successive differences (RMSSD) (1,2). Skin temperature (ST) was recorded for every 1 minute that the device was worn; these data were filtered to only those recordings that occurred during sleep (3). Respiratory rate (RR) was provided only as an average rather than a time series.

For HR, HRV, and ST, three candidate methods of summarizing each set of nightly measurements into one nightly summary value were considered, i.e., taking the 1st percentile, the mean, and the 99th percentile. For a given evening and domain, these nightly summary values were only calculated where the domain was successfully recorded for the whole sleep period.

For each of the 10 nightly summary variables (3 for HR, 3 for HRV, 3 for ST, and 1 for RR), a subject norm was calculated for each subject by averaging over the available measurements across a 10-night period from nights 7 through 16 after dose one. The start of this window was chosen because vaccination side effects have generally resolved by this time (4,5), and no subjects received a second dose by night 16. Nightly summary variables were then centered on each subject norm.

Nightly statistics were then collapsed across nights to generate per-subject summary statistics of vaccination-induced change. For nights 0 and 1 following each dose, the nightly mean HR and ST was positive on average, while for HRV it was negative on average, indicating that the direction of any effect of vaccination on HR, RR, and ST was an increase, while for HRV it was a decrease. Because subjects received vaccination at varying times of day, between-subject variability in the time to maximal vaccination-induced change was anticipated. Across nights 0 through 3 following each dose, the majority of subjects experienced their peak deviation from baseline (maximum for mean nightly HR, RR, and ST; minimum for mean nightly HRV) on either night 0 or 1, so the larger deviation over these two nights was taken as each subject’s vaccination-induced change in each of the ten variables, for each vaccine dose.

The nightly summary method (i.e., 1^st^ percentile, mean, or 99^th^ percentile) most sensitive to vaccination-induced change in each of HR, HRV, and ST was then identified by using one sample t-tests to compare the dose two summary variables derived from the nightly 1^st^ percentile, mean, and 99^th^ percentile values. Dose two was used for the purpose of variable selection because side effects are more common at dose two and therefore variable comparison was less likely to be influenced by random variability (5). The most sensitive variable was defined as the one with the lowest p-value. Results are provided in Supplement Table 2. The nightly 99^th^ percentile was identified as most sensitive to vaccination-induced change for HRV and ST, while the nightly 1^st^ percentile was most sensitive to vaccination-induced change for HR.

### Data analysis

Linear mixed-effects models were fit using *lme4* using restricted maximum likelihood estimation. Deviation (sum-to-zero) coding was used for categorical predictors. Continuous predictors were mean centered prior to model fitting. Random effects included a random intercept per subject. All models were checked for multicollinearity by examination of variance inflation factors (VIFs) calculated using *car*. The VIF threshold of 5 was exceeded in the initial model for each of the following predictors of interest: other allergic reactions (dose one), pain or swelling in the non-injected arm (dose one), tender or swollen lymph nodes (dose one), fever (dose one), other allergic reactions (doses one and two). In all cases, multicollinearity was the result of low variability in these predictors, resulting in the higher-order two- or three-way interaction terms being correlated with vaccine, time point, or both. This was resolved by removing all interaction terms involving each predictor of interest. All final models met assumptions of residual normality, linearity, and equality of variance, as assessed via diagnostic plots. No single observation had an undue influence on model fit, given that all had a Cook’s distance below 1. Significance of model terms was evaluated via F statistics calculated using Type II sums of squares, meaning that the F statistic corresponding to a given term compared the predictions of the full model including that term but without any higher-order interaction terms to the same model without the given term (6). Degrees of freedom were approximated via the Kenward-Roger method.

Along with test statistics, results were described using marginal means (i.e., least-squares means) or using unstandardized or standardized effect size estimates. Unstandardized effect sizes included marginal slopes or the difference between a pair of marginal means or slopes, both calculated using *emmeans*. The *effectsize* package was used to convert t or Type II sums of squares F statistics and associated degrees of freedom to standardized effect sizes, specifically the partial correlation (r_p_) or partial η2. Effect sizes were also presented as fold difference where appropriate. All estimates assumed (i.e., were conditioned on) mean levels of continuous covariates (age and body mass index) and were averaged across the estimates for each level of other categorical predictors (vaccine, sex, baseline smoking status, time point), weighting each level of these variables proportional to its representation in the sample. Post-hoc testing was performed on these estimates, comparing two using a two-sample t-test, or comparing marginal slopes (simple slopes) to zero using a one-sample t-test. Marginal means were provided in the untransformed (ID50) scale, and fold differences were calculated in this scale. Marginal trends and pairwise contrasts refer to associations with the outcome in the log10 scale. All visualizations were produced using *ggplot2* and *patchwork*, with *ggeffects* used for calculation of partial residuals.

### Supplement Figures

**Supplement Figure 1.**
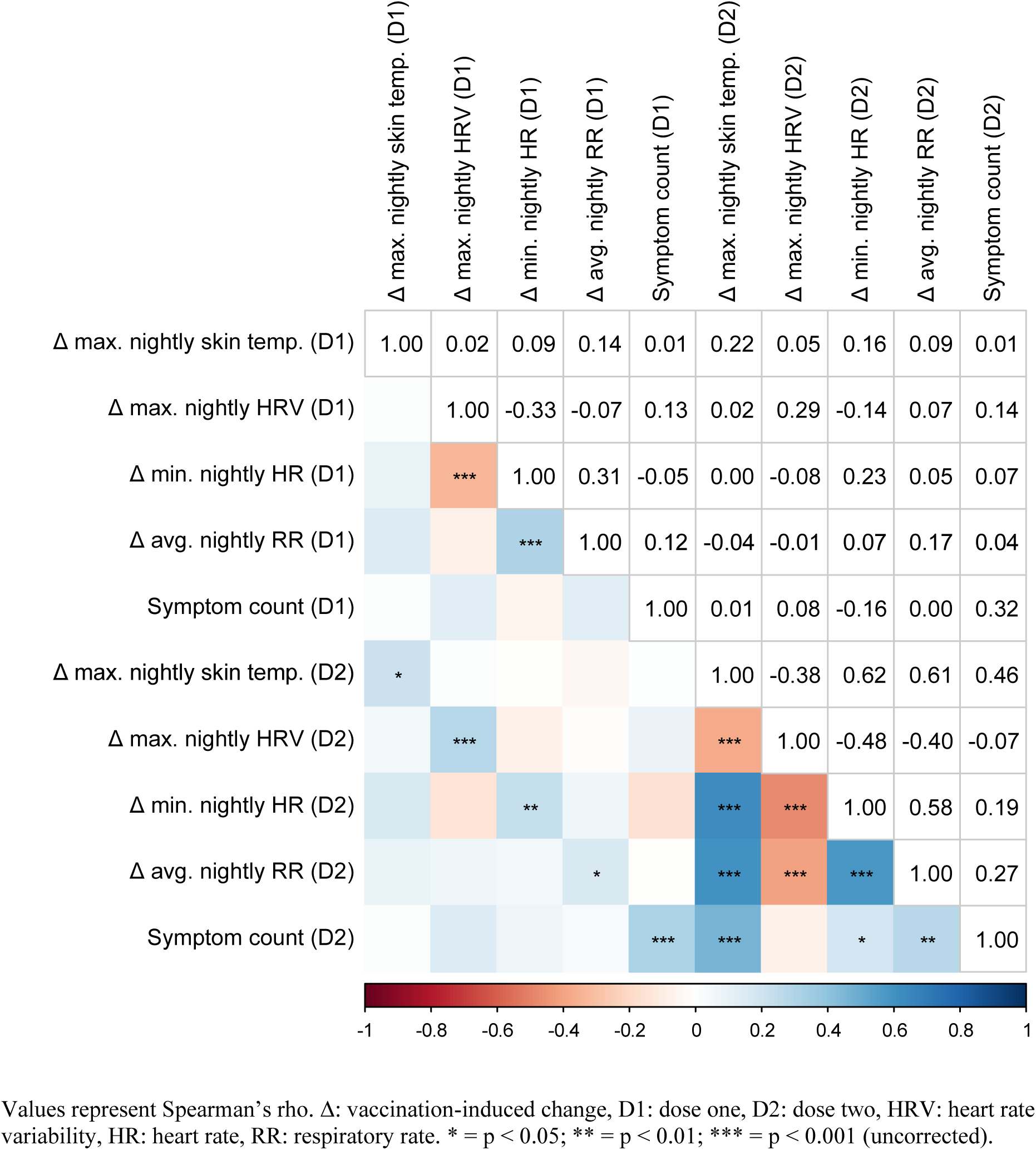
Correlations between continuous predictor variables. Values represent Spearman’s rho. Δ: vaccination-induced change, D1: dose one, D2: dose two, HRV: heart rate variability, HR: heart rate, RR: respiratory rate. * = p < 0.05; ** = p < 0.01; *** = p < 0.001 (uncorrected).

## References

1. Wu N, et al. Long-term effectiveness of COVID-19 vaccines against infections, hospitalisations, and mortality in adults: findings from a rapid living systematic evidence synthesis and meta-analysis up to December, 2022. Lancet Respir Med. 2023;11(5):439–52. doi:10.1016/S2213-2600(23)00015-2

2. Centers for Disease Control and Prevention. COVID Data Tracker [Internet]. Atlanta, GA: U.S. Department of Health and Human Services, CDC. 2023 [updated 2023 Sep 21; cited 2023 Sep 21].

3. Sinclair AH, Taylor MK, Weitz JS, Beckett SJ, Samanez-Larkin GR. Reasons for Receiving or Not Receiving Bivalent COVID-19 Booster Vaccinations Among Adults — United States, November 1–December 10, 2022. MMWR Morb Mortal Wkly Rep. 2023;72(3):72–5. doi:10.15585/mmwr.mm7203a5

4. Jacobs ET, et al. Understanding low COVID-19 booster uptake among US adults. Vaccine. 2023. doi:10.1016/j.vaccine.2023.08.080

5. Hermann EA, et al. Association of Symptoms After COVID-19 Vaccination With Anti-SARS-CoV-2 Antibody Response in the Framingham Heart Study. JAMA Netw Open. 2022;5(10):e2237908. doi:10.1001/jamanetworkopen.2022.37908

6. Debes AK, et al. Association of Vaccine Type and Prior SARS-CoV-2 Infection With Symptoms and Antibody Measurements Following Vaccination Among Health Care Workers. JAMA Intern Med. 2021;181(12):1660–2. doi:10.1001/jamainternmed.2021.4580

7. Tani N, et al. Relation of fever intensity and antipyretic use with specific antibody response after two doses of the BNT162b2 mRNA vaccine. Vaccine. 2022;40(13):2062–7. doi:10.1016/j.vaccine.2022.02.025

8. Choi MJ, et al. Predictive Value of Reactogenicity for Anti-SARS-CoV-2 Antibody Response in mRNA-1273 Recipients: A Multicenter Prospective Cohort Study. Vaccines (Basel). 2023;11(1). doi:10.3390/vaccines11010120

9. Yoshida M, et al. Association of systemic adverse reaction patterns with long-term dynamics of humoral and cellular immunity after coronavirus disease 2019 third vaccination. Sci Rep. 2023;13(1):9264. doi:10.1038/s41598-023-36429-1

10. Moncunill G, et al. Determinants of early antibody responses to COVID-19 mRNA vaccines in a cohort of exposed and naïve healthcare workers. EBioMedicine. 2022;75103805. doi:10.1016/j.ebiom.2021.103805

11. Levin EG, et al. Waning Immune Humoral Response to BNT162b2 Covid-19 Vaccine over 6 Months. N Engl J Med. 2021;385(24):e84. doi:10.1056/NEJMoa2114583

12. Gilbert PB, et al. Immune correlates analysis of the mRNA-1273 COVID-19 vaccine efficacy clinical trial. Science. 2022;375(6576):43–50. doi:10.1126/science.abm3425

13. Rogers TF, et al. Isolation of potent SARS-CoV-2 neutralizing antibodies and protection from disease in a small animal model. Science. 2020;369(6506):956–63. doi:10.1126/science.abc7520

14. Cromer D, et al. Neutralising antibody titres as predictors of protection against SARS-CoV-2 variants and the impact of boosting: a meta-analysis. Lancet Microbe. 2022;3(1):e52–e61. doi:10.1016/S2666-5247(21)00267-6

15. Khoury DS, et al. Neutralizing antibody levels are highly predictive of immune protection from symptomatic SARS-CoV-2 infection. Nat Med. 2021;27(7):1205–11. doi:10.1038/s41591-021-01377-8

16. Prather AA, et al. Predictors of long-term neutralizing antibody titers following COVID-19 vaccination by three vaccine types: the BOOST study. Sci Rep. 2023;13(1):6505. doi:10.1038/s41598-023-33320-x

17. Shilaih M, Goodale BM, Falco L, Kübler F, Clerck V de, Leeners B. Modern fertility awareness methods: wrist wearables capture the changes in temperature associated with the menstrual cycle. Biosci Rep. 2018;38(6). doi:10.1042/BSR20171279

18. Zhu TY, et al. The Accuracy of Wrist Skin Temperature in Detecting Ovulation Compared to Basal Body Temperature: Prospective Comparative Diagnostic Accuracy Study. J Med Internet Res. 2021;23(6):e20710. doi:10.2196/20710

19. Kung Y-A, et al. Factors influencing neutralizing antibody titers elicited by coronavirus disease 2019 vaccines. Microbes Infect. 2023;25(1-2):105044. doi:10.1016/j.micinf.2022.105044

20. Dieckhaus KD, et al. SARS-CoV-2 Antibody Dynamics in Healthcare Workers after mRNA Vaccination. Vaccines (Basel). 2023;11(2). doi:10.3390/vaccines11020358

21. Cheng A, et al. Correlation of adverse effects and antibody responses following homologous and heterologous COVID19 prime-boost vaccinations. J Formos Med Assoc. 2023;122(5):384–92. doi:10.1016/j.jfma.2022.12.002

22. Mason AE, et al. Metrics from Wearable Devices as Candidate Predictors of Antibody Response Following Vaccination against COVID-19: Data from the Second TemPredict Study. Vaccines (Basel). 2022;10(2). doi:10.3390/vaccines10020264

23. Zhong D, et al. Durability of Antibody Levels After Vaccination With mRNA SARS-CoV-2 Vaccine in Individuals With or Without Prior Infection. JAMA. 2021;326(24):2524–6. doi:10.1001/jama.2021.19996

24. Menni C, et al. Vaccine side-effects and SARS-CoV-2 infection after vaccination in users of the COVID Symptom Study app in the UK: a prospective observational study. Lancet Infect Dis. 2021;21(7):939–49. doi:10.1016/S1473-3099(21)00224-3

25. Sette A, Sidney J, Crotty S. T Cell Responses to SARS-CoV-2. Annu Rev Immunol. 2023;41343–73. doi:10.1146/annurev-immunol-101721-061120

26. Bertoletti A, Le Bert N, Tan AT. SARS-CoV-2-specific T cells in the changing landscape of the COVID-19 pandemic. Immunity. 2022;55(10):1764–78. doi:10.1016/j.immuni.2022.08.008

## Supplement References

1. Kinnunen H, Rantanen A, Kenttä T, Koskimäki H. Feasible assessment of recovery and cardiovascular health: accuracy of nocturnal HR and HRV assessed via ring PPG in comparison to medical grade ECG. Physiol Meas. 2020;41(4):04NT01. doi:10.1088/1361-6579/ab840a

2. Cao R, et al. Accuracy Assessment of Oura Ring Nocturnal Heart Rate and Heart Rate Variability in Comparison With Electrocardiography in Time and Frequency Domains: Comprehensive Analysis. J Med Internet Res. 2022;24(1):e27487. doi:10.2196/27487

3. Maijala A, Kinnunen H, Koskimäki H, Jämsä T, Kangas M. Nocturnal finger skin temperature in menstrual cycle tracking: ambulatory pilot study using a wearable Oura ring. BMC Womens Health. 2019;19(1):150. doi:10.1186/s12905-019-0844-9

4. Menni C, et al. Vaccine side-effects and SARS-CoV-2 infection after vaccination in users of the COVID Symptom Study app in the UK: a prospective observational study. Lancet Infect Dis. 2021;21(7):939–49. doi:10.1016/S1473-3099(21)00224-3

5. Rosenblum HG, et al. Safety of mRNA vaccines administered during the initial 6 months of the US COVID-19 vaccination programme: an observational study of reports to the Vaccine Adverse Event Reporting System and v-safe. Lancet Infect Dis. 2022;22(6):802–12. doi:10.1016/S1473-3099(22)00054-8

6. Langsrud Ø. ANOVA for unbalanced data: Use Type II instead of Type III sums of squares. Statistics and Computing. 2003;13(2):163–7. doi:10.1023/A:1023260610025

